# Mapping the Bird Risk Index for West Nile virus in Europe and its relationship with disease occurrence in humans

**DOI:** 10.1101/2025.02.24.25322781

**Authors:** Jonathan Bastard, Raphaëlle Métras, Benoit Durand

## Abstract

West Nile virus (WNV) is a zoonotic mosquito-borne *Flavivirus*, with bird populations reservoirs. Although often asymptomatic, infection in humans can cause febrile symptoms and, more rarely, severe neurological symptoms. Previous studies assessed environmental drivers of WNV infections, but most relied on notified West Nile Disease human cases, exposing them to (i) overlook areas with WNV circulation despite no reported case, and (ii) mixing mechanisms affecting hosts *vs.* vectors. Our objective was to generate a WNV Bird Risk Index (BRI), mapping the potential of WNV circulation in bird communities across Europe, in order to better understand the distribution of WNV infections. We first used a bird traits-based model to estimate WNV seroprevalence in European bird species. This allowed us to build a map of the WNV BRI across Europe. To validate this metric, we investigated its association with WNV human cases notified at the NUTS administrative region scale, using a Besag-York-Mollie 2 spatial model in a Bayesian framework. We first identified eco-ethological characteristics associated with higher WNV seroprevalence in wild birds. Second, we mapped the BRI that showed a strong spatial heterogeneity across Europe. At the NUTS level, the BRI was positively associated with the number of years with notified WNV human cases between 2016 and 2023. To conclude, we provide a map quantifying the suitability for WNV to circulate in the bird reservoir. This allows to target surveillance efforts in areas at risk for WNV zoonotic infections in the future.

## Introduction

The occurrence of arboviral diseases in humans either results from an epidemiological cycle involving mostly humans (such as dengue or chikungunya, although sylvatic transmissions have also been reported for these viruses [1]), zoonotic transmissions from an animal reservoir source (such as West Nile or tick-borne encephalitis), or a combination of both (such as yellow fever). In the zoonotic case, the risk of infection to humans may result from a combination of (i) the level of infection in the animal reservoir – depending on the structure of both the host and vector community – that represents a zoonotic *potential*, thereafter named the structural risk, and (ii) environmental (e.g. high mosquito abundance following rains) or anthropic (e.g. control measures against mosquitoes) circumstances that favour subsequent zoonotic transmission events to humans, thereafter named the conjectural risk [2]. The structural risk is typically expected to change on the longer term than the conjectural risk, because the former depends on the global spatial distribution of species that changes at the scale of several years or decades [3], as compared to the week or month scale for the latter.

West Nile virus (WNV) is a mosquito-borne *Flavivirus* with bird populations as reservoirs and *Culex* mosquitoes as main vector [4]. It can also infect humans and horses, considered as “dead-end” hosts because they do not transmit further the virus to biting mosquitoes. However, human-to-human transmission remains possible through the donation of blood or organs from an infected donor [5]. Although asymptomatic in about 80% of infections in humans, WNV can cause febrile symptoms and, in rare cases, severe neurological symptoms that may lead to death [4]. For these reasons, mapping WNV infection risk, despite its possible silent circulation, is important to strengthen surveillance and preparedness in affected areas.

Previous studies investigated spatial drivers of WNV infections at the continental scale of Europe, North America or Africa [2,6–15]. Most of them assessed environmental factors associated with West Nile disease (WND) cases reported in humans or animals, which included temperature, precipitations, land type and vegetation cover, among others [8,9,11–15]. However, this approach that can be described as a data-driven or cases-to-reservoir approach, may pose two types of issues.

First, relying on reports of WNV infection in humans, animals or vectors to fit spatial models and derive environmental drivers may not allow to distinguish between a true absence of viral circulation in the reservoir and an absence of reported infections (e.g. in areas with lower human or equine exposure, or with lower surveillance efforts).

Second, it does not discriminate the effects of environment on the reservoir host community (associated to the structural risk) *versus* on the abundance of bridge vectors (also associated to the conjectural risk), leading to conflicting statistical associations. For instance, local temperature patterns may affect both the suitability for key reservoir bird species and the dynamics of *Culex* during the mosquito season, which both ultimately influence human WND occurrence. This may be why the associations between WND case occurrence and some environmental variables depends on the season, for example the temperature in winter *versus* in summer [11] or the relative humidity in spring *versus* in autumn [12].

Instead, separating these components of human infection risk as [2,10] allows (i) to better understand WNV epidemiological system, and (ii) to adapt monitoring and control strategies focusing on long-term (e.g. implementing a surveillance scheme) or short-term (e.g. timely increasing awareness of the public or practitioners) measures. Here, we adopted this approach that can be described as knowledge-driven, or reservoir-to-cases, because it is based on assumptions about the eco-epidemiological mechanisms driving the circulation of this zoonotic pathogen.

We first quantified the virus’ structural risk across Europe using recent data on bird host spatial distribution and WNV seroprevalence, producing a WNV Bird Risk Index (BRI) that showed spatial heterogeneity across Europe. We then validated this measure against long-term trends of WND occurrence in humans over eight years.

## Materials and Methods

### Review of serological surveys in birds

We reviewed the literature to find WNV serological data on European bird species (step A in Figure 1). First, we retrieved articles included in a previous literature review published in 2017 [2]. Second, on March 6^th^ 2024, we searched the following terms on PubMed: “(West Nile [title]) AND (bird* [title/abstract] OR avian [title/abstract]) AND (sero* [title/abstract]) AND ("2016/01/01" [Date - Publication]: "3000" [Date - Publication]) NOT (Review[Publication Type])”. Inclusion criteria were: articles written in English or French, samples in European birds weighting less than 50g (see list of the 150 species in Supplementary Data 1), information available on the number of both negative and positive samples, confirmation of positive samples by virus neutralisation tests. We focused on birds weighting less than 50g because they tend to have shorter lifespans (a few years) than larger birds [16,17], and thus a higher proportion of susceptible individuals (due to population renewal) for any given force of infection and virus-induced lethality. Moreover, lighter birds tend to be more abundant than heavier birds, as suggested by the literature [18] and by comparing the relative abundances provided by eBird [19,20] for a subset of 65 European species weighting less than 50g (median cumulated relative abundance of 25.4, as defined in the legend of Supplementary Figure S1, across the study area) as compared to *Corvidae* (median of 2.1 for 5 species) and raptor species (median of 1.5 for 36 species) (Supplementary Figure S1). These points lead to expect (i) a substantial role of smaller birds in WNV epidemiological cycle [2,21], although it also depends on host competence that has been little studied for European species (e.g. [22,23]), and (ii) that the seroprevalence in these bird species allows to better assess the force of infection applying to them [24].

**Figure 1.**
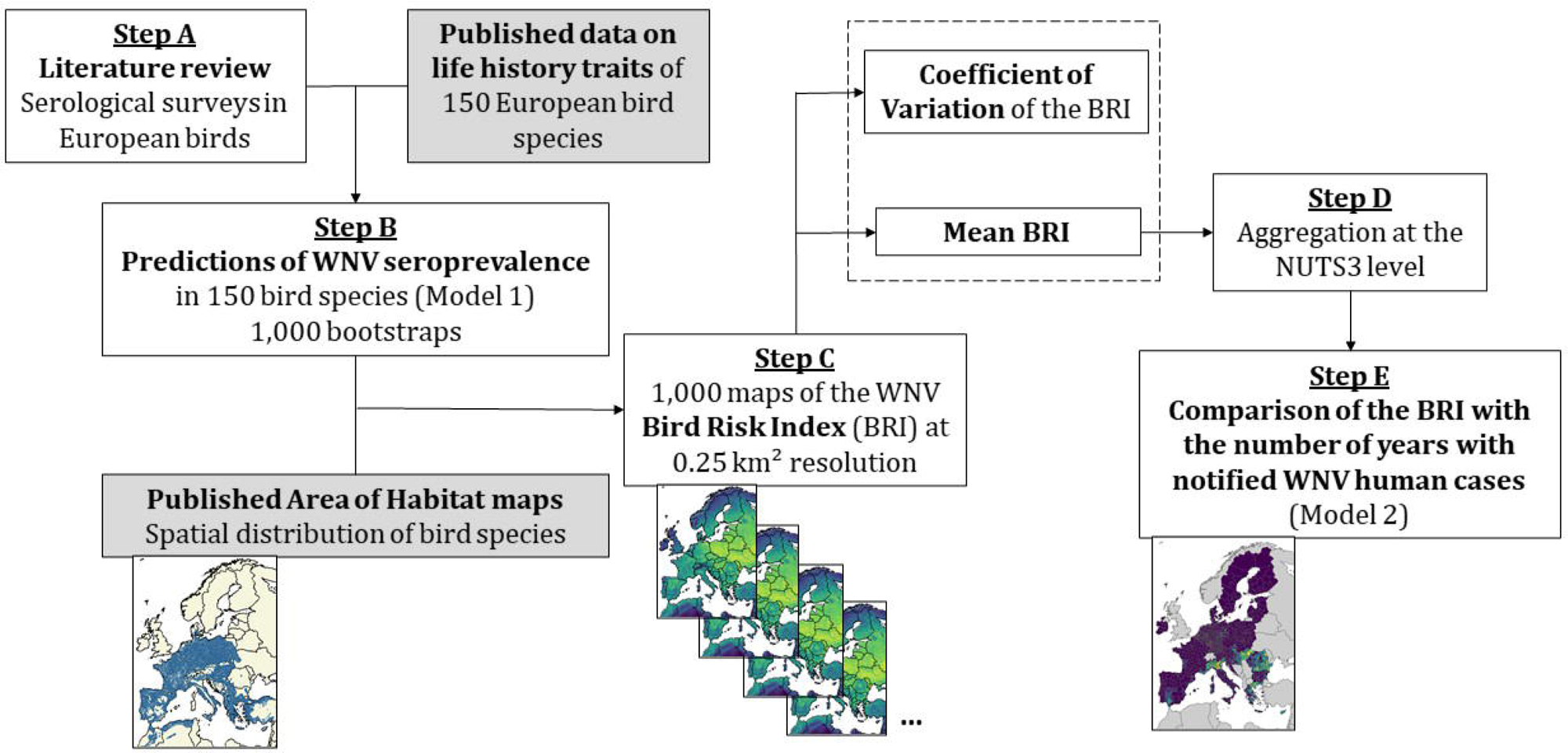
Diagram of the methodology to map West Nile Virus (WNV) Bird Risk Index (BRI) and validate it against WNV human cases data. The white boxes represent the steps of the analyses presented in this paper, whilst grey boxes show input information used from published studies. We first performed a literature review of WNV seroprevalence surveys in European bird species (step A). Using this seroprevalence data and previously published species traits information [2], we predicted WNV seroprevalence in 150 bird species using a mixed-effect logistic regression with 1,000 bootstrap replicates (step B). We then combined each of the 1,000 bootstrap results to published distribution maps for the 150 bird species [25] to compute 1,000 maps of the BRI in Europe at a 0.25 km² resolution (step C). For each pixel, we calculated the mean (averaged over 1,000 replicates) and Coefficient of Variation of the BRI, and aggregated them at the NUTS administrative level (step D). We finally used the mean BRI as predictor in a spatial model predicting the number of years with notified WNV human cases between 2016 and 2023 (step E).

### Predictions of seroprevalence in birds

Using the values of seroprevalence found in the literature for some bird species, we aimed at inferring seroprevalence estimates for 150 European bird species weighting less than 50g (step B in Figure 1). We fitted a mixed-effects logistic model where the outcome was the serological status (negative or positive) of individuals sampled in the studies (hereafter named Model 1). Predictors included as fixed effects were life history traits determined for each of the 150 bird species by expert opinion and literature search, as described in a previously published study [2] (Supplementary Data 1), namely the migratory status (yes/no), body mass, nest height (ground, intermediate or above 4 metres), use of suburban or urban habitats (yes/no), nocturnal gregariousness (yes/no), exposure of nestlings (altricial/precocial) and breeding sociality (yes/no). The ecological justifications for including each of these trait variables are provided in Supplementary Table S1. Study identification numbers were included as random effect to account for the variability of sampling years and locations in the literature. Confidence intervals for the seroprevalence predicted in bird species were generated by bootstrapping using 1,000 replicates.

### Spatial distribution of the West Nile Bird Risk Index

To obtain the spatial distribution of West Nile virus exposure potential in birds, we then combined the seroprevalence predictions in the 150 bird species to published maps of their spatial distribution [25] (step C in Figure 1). Briefly, Lumbierres et al. produced presence/absence habitat maps at 0.01 km² resolution by subtracting areas deemed unsuitable for the species (based on their land cover and elevation preferences) from their global range maps. Instead, we worked at the 0.25 km² resolution for computational efficiency purposes by considering *P_kj_*, the proportion of 0.01 km² pixels where species j was present within each 0.25 km² pixel *k*. The Bird Risk Index (BRI) was then defined as:

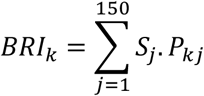

Where *S_j_* was WNV seroprevalence predicted in species *j* by Model 1. We generated 1,000 BRI maps from the 1,000 bootstrap replicates, in each of which BRI values were standardised between 0 and 1. We subsequently produced maps of the mean and coefficient of variation values of the BRI.

### Prediction of West Nile disease occurrence in Europe

We then assessed the relationship between BRI and a time-aggregated outcome representing WN human disease risk in the European Union (steps D and E in Figure 1), namely the number of years with human cases notified at the NUTS administrative region level between 2016 and 2023 (Model 2). The case report data was obtained from the European Centre for Disease Prevention and Control [26]. We performed analyses at the NUTS3 level, excepted in countries where the average surface area of NUTS2 regions was closer to the European average NUTS3 surface area. This was the case for Belgium, Germany, Malta and Netherlands, where cases reported at the NUTS3 level were aggregated at the NUTS2 level.

We assumed a binomial distribution for Model 2, with 8 trials corresponding to a maximum of 8 years with WND cases in the data. The mean BRI used as fixed effect was averaged over all pixels of each NUTS region. We also included the log-scaled population density, obtained from Eurostat [27], as a fixed effect in Model 2. We added a country random effect to account for variations in surveillance exhaustiveness and other national socioeconomic or cultural factors affecting the occurrence and notification of human cases. In addition, since some countries contained only one NUTS region, we combined those with neighbouring countries regarding the country effect, to avoid convergence issues at the model fitting stage. Malta was combined with Italy, Luxembourg with Belgium and Cyprus with Greece. We also accounted for spatial autocorrelation of WND occurrence in NUTS regions using a Besag-York-Mollie 2 (BYM2) model, which combines spatially structured and unstructured effects [28]. We fitted the models with the R-INLA package, version 23.04.24 [29].

We performed a 5-fold cross-validation of the model, where the sampling of folds was stratified by country and presence of cases in the NUTS region. In each case, we fitted the model to the training dataset and assessed the deviance-based R², as defined by [30] for count data, to compare model predictions and the testing data. We considered both the overall R² (i.e. the conditional R² accounting for both fixed and random effects) and the marginal R² (accounting for the fixed effect only). We also computed the Probability Integral Transform (PIT) histograms by implementing a non-randomised method for count data described by [31,32]. When the consistency between observations and model predictions is good, the PIT histogram approximately follows a uniform distribution. We included all the data in the final model and assessed its stability by checking that coefficients’ posterior estimates and hyperparameters of the random effects were then similar to values from the training data only. Furthermore, we compared the final model with a null model, both with and without the random effect, using the Deviance Information Criterion (DIC).

### Sensitivity analysis

We first evaluated the sensitivity of our results to the predictor applied in Model 2. We either removed the “mean BRI” fixed effect, or replaced it by (i) the species richness, i.e. a variant of the Bird Risk Index not considering variability in seroprevalence values across bird species, or (ii) the proportion of bootstrap replicates for which each pixel fell in the 50% highest pixel values, hereafter named P50. We computed the DIC for these substitute models and compared them to the main analysis. Second, we fitted an alternative model, named Model 2bis, considering as outcome the cumulated number of notified human cases between 2016 and 2023, with a negative binomial distribution, the mean BRI as a fixed effect, the log-scaled population density as an offset, and the same random effects as above. We checked the goodness of fit for this alternative as described above.

## Results

### Review of serological surveys in birds

Twenty-nine studies corresponded to inclusion criteria (see diagram flow and references in Supplementary Figure S2), reporting 11,989 sera samples in 73 bird species. The highest number of samples was in *Passer domesticus* and *Hirundo rustica* (n=5,999 and 1,335 samples respectively). Globally, 1.6% (192/11989) of published sample results were positive, although the proportion depended on the species as detailed in Supplementary Data 2.

### Predictions of WNV seroprevalence in birds

Using published serological results and data on life history traits, we predicted WNV seroprevalence in European birds weighting less than 50g. Among 150 bird species, point predictions ranged between 0.48% (for *Riparia riparia*) and 9.67% (for *Phylloscopus borealis*), although with a large variability of bootstrap confidence interval sizes (Supplementary Figure S3).

WNV seroprevalence was found to be higher in altricial species than in precocial species (odds-ratio (OR) at 2.15 with a 95% confidence interval of [1.38; 3.35]), and lower for species showing nocturnal gregariousness (OR = 0.61, 95% CI [0.38; 0.97]). Species nesting on the ground were found more WNV seropositive (OR = 2.1, 95%CI [1.18; 3.74]) than species nesting above ground (Table 1). On the contrary, no significant effect was found for the migratory status, body mass (under 50g), use of urban or suburban habitat and breeding sociality (Table 1).

**Table 1.**
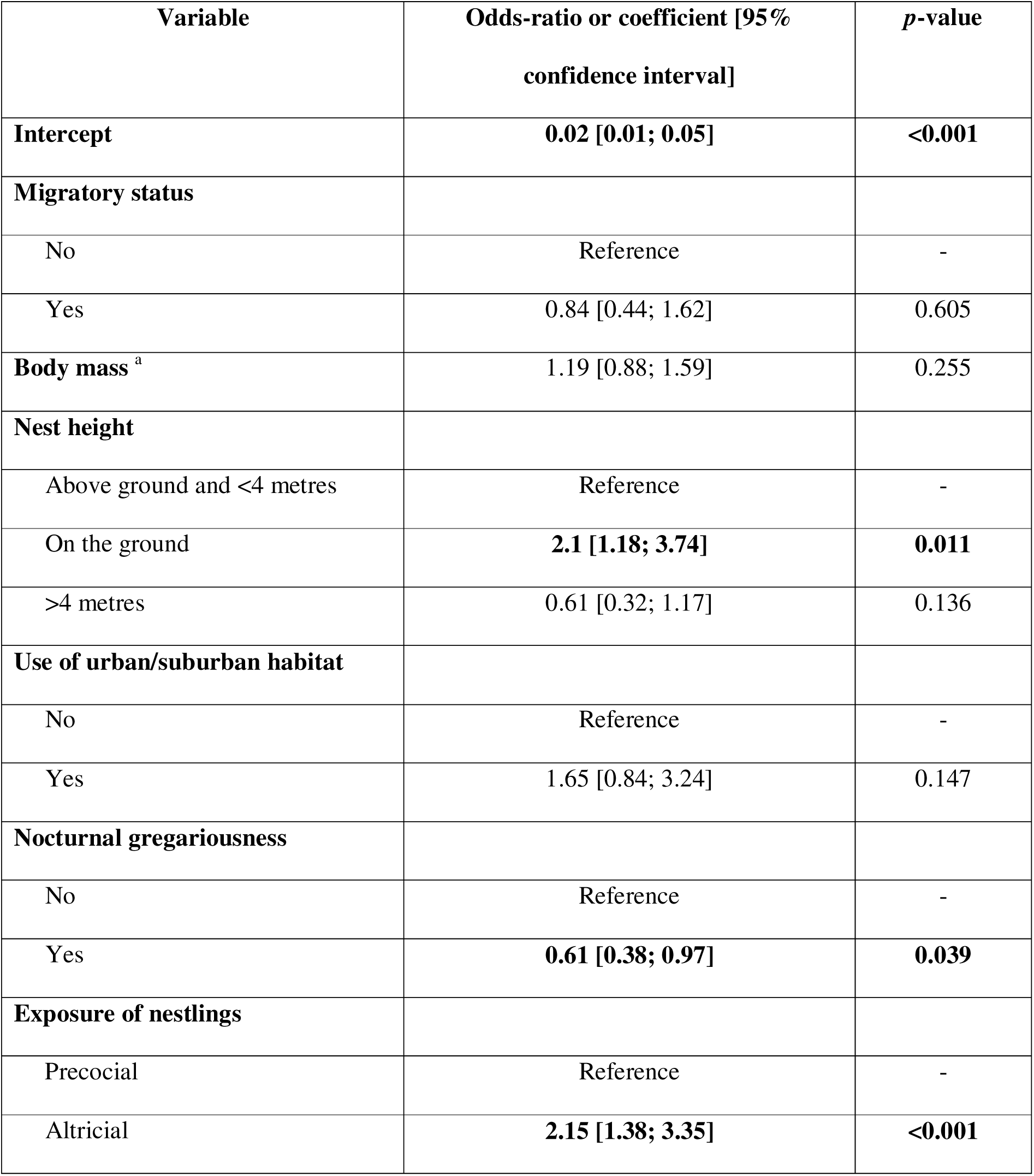
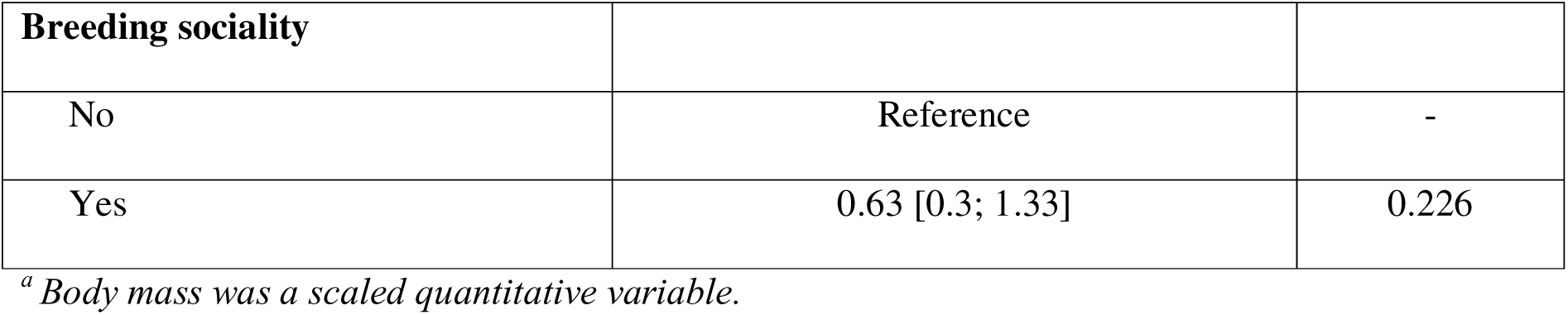
Odds-ratios or coefficients of the mixed-effects logistic model predicting seroprevalence in bird species (Model 1).

| Variable | Odds-ratio or coefficient [95% confidence interval] | *p*-value |
| --- | --- | --- |
| **Intercept** | **0.02 [0.01; 0.05]** | **<0.001** |
| **Migratory status** |  |  |
| No | Reference | - |
| Yes | 0.84 [0.44; 1.62] | 0.605 |
| **Body mass<sup>a</sup>** | 1.19 [0.88; 1.59] | 0.255 |
| **Nest height** |  |  |
| Above ground and <4 metres | Reference | - |
| On the ground | **2.1 [1.18; 3.74]** | **0.011** |
| >4 metres | 0.61 [0.32; 1.17] | 0.136 |
| **Use of urban/suburban habitat** |  |  |
| No | Reference | - |
| Yes | 1.65 [0.84; 3.24] | 0.147 |
| **Nocturnal gregariousness** |  |  |
| No | Reference | - |
| Yes | **0.61 [0.38; 0.97]** | **0.039** |
| **Exposure of nestlings** |  |  |
| Precocial | Reference | - |
| Altricial | **2.15 [1.38; 3.35]** | **<0.001** |
| **Breeding sociality** |  |  |
| No | Reference | - |
| Yes | 0.63 [0.3; 1.33] | 0.226 |

<sup>a</sup> *Body mass was a scaled quantitative variable.*

### Relationship between the BRI and West Nile disease occurrence in Europe

Figure 2 depicts the spatial distribution of West Nile BRI computed in Europe and around the Mediterranean basin. Mean BRI values across pixels ranged between 0 and 0.98, with a median of 0.45. Areas with higher values (closer to 1) may be interpreted as showing more potential (or likelihood) for WNV circulation in the birds community. While varying at small geographical scale, the reservoir potential was overall higher on the Central and Eastern parts than the Western and Northern parts of the continent. Highest uncertainty was found in the northernmost parts of Europe and South to the Atlas Mountains (Figure 2).

**Figure 2.**
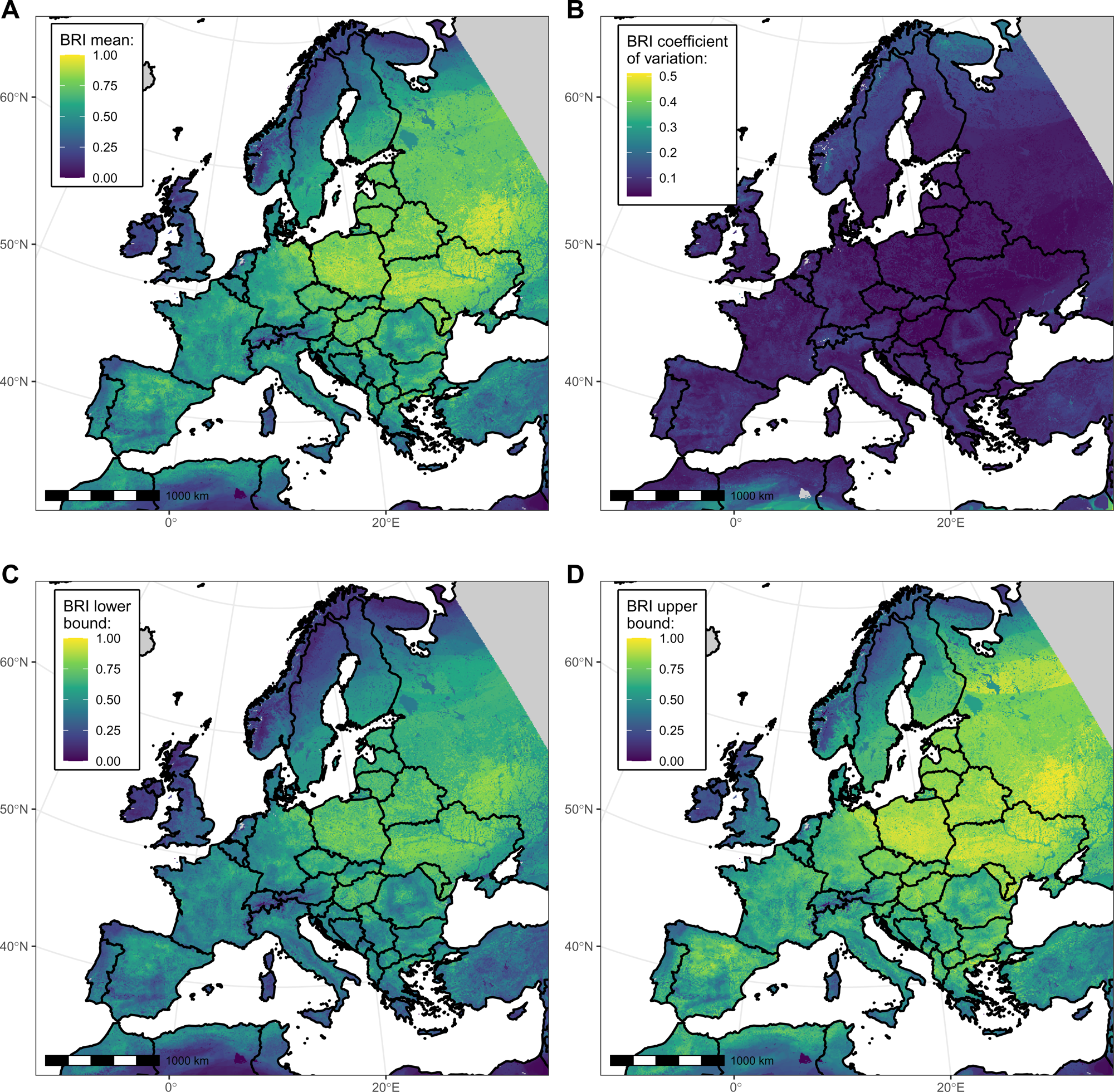
Maps showing West Nile Virus Bird Risk Index (BRI) mean (panel A), coefficient of variation (panel B), and lower and upper bounds of the 95% confidence interval (panels C and D), across bootstrap replicates at the 0.25 km² resolution. Areas with no value are shown in grey. For computational efficiency reasons, only a subset of 200 bootstrap replicates were used to derive the confidence interval bounds.

We then assessed the association between the BRI and WND human cases notified at the NUTS administrative region scale (Model 2). During cross-validation, Model 2 predictions provided satisfying consistency with the testing data (pooled overall and marginal R² values of 0.57 and 0.28 respectively), as shown in Supplementary Table S2 and in PIT histograms (Supplementary Figure S4). Hyperparameters of the BYM2 and “Country” random effects were stable (pooled BYM2 precision of 0.64). The goodness of fit of the final model was also satisfying, as suggested by the PIT histogram (Supplementary Figure S5). Deviance residuals of the model are shown in Supplementary Figure S6. The DIC of the final model was lower than the null model, both with and without random effect (Supplementary Table S3). Model 2 correctly predicted spatial clusters of notified cases in Northern Italy, Northern Greece, Hungary, Romania and Eastern Germany (Figure 3).

**Figure 3.**
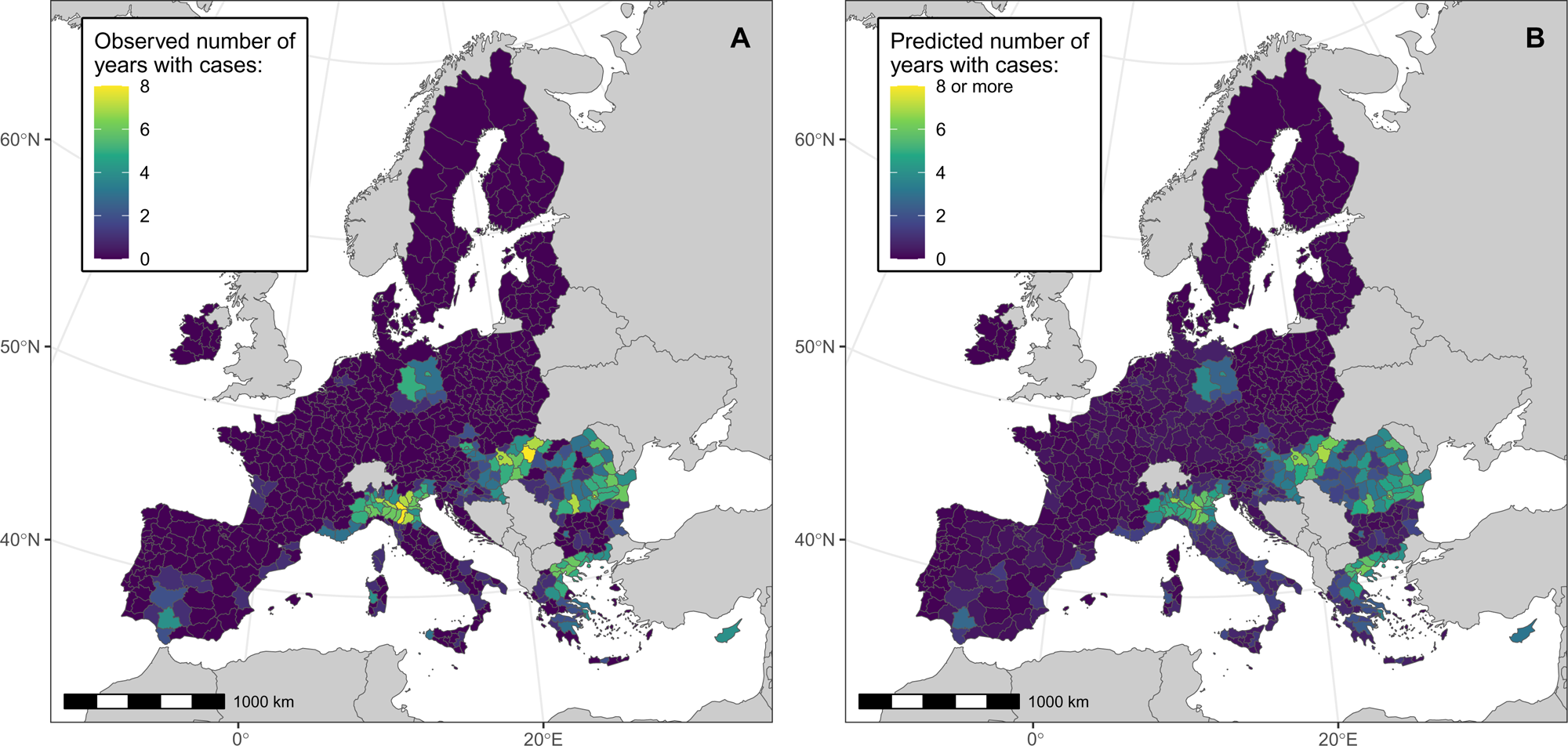
Model 2 maps showing the observed (panel A) and predicted (panel B) number of years with notified WNV human cases in NUTS European administrative regions, between 2016 and 2023. We performed analyses at the NUTS3 level, or NUTS2 level for Belgium, Germany, Malta and Netherlands.

At the NUTS level, the mean BRI was positively associated with the number of years with notified WND cases (Table 2). Posterior estimates were similar across cross-validation folds (Supplementary Table S2).

**Table 2.** Results of Model 2, which predicts the number of years with notified WNV human cases in NUTS European administrative regions between 2016 and 2023. Coefficient values are presented with their 95% posterior interval.

| Variable | Coefficient posterior median [95% credible interval] |
| --- | --- |
| Intercept | -24.7 [-29.2; -20.6] |
| Mean BRI | 9.97 [7.82; 12.2] |
| log(population density) | 1.06 [0.80; 1.32] |

*BRI: Bird Risk Index.*

### Sensitivity analysis

In Model 2, removing the “mean BRI” variable or changing it to the species richness or P50 led to higher DIC values than the baseline analysis (Supplementary Table S3), hence could not be considered as improving the model. Similar to Model 2, the alternative Model 2bis exhibited a positive association between the BRI and the cumulated number of notified WND cases between 2016 and 2023 at the NUTS level (Supplementary Table S4). Moreover, risk areas predicted by Model 2bis seemed globally consistent with observations and Model 2 predictions (Supplementary Figure S7). However, for Model 2bis, PIT histograms suggested disparities between observed and predicted distributions at the cross-validation step (Supplementary Figure S8) and when the full dataset was used (Supplementary Figure S5). This calls for caution when interpreting Model 2bis results.

## Discussion

In this study, we built a map of the potential of birds’ exposure to West Nile virus (WNV), the Bird Risk Index (BRI), quantifying the spatial distribution of the virus’ structural risk across Europe. This metric was positively associated to the occurrence of notified West Nile human cases at the NUTS administrative level.

Our results were partly in line with previously published risk maps of WNV in Europe, with overall higher BRI values east than west of the continent similarly to [7–9,11,12]. However, we identified areas that were discordant with some of the previous studies based on reported WNV human cases, such as Poland and Baltic countries for instance [7,11,12]. These discrepancies may first be explained by a circulation of WNV in wild birds that translates into no or few zoonotic infections due to a low human exposure related to local landscape, anthropic (population distribution, control measures against mosquitoes, etc.) or vector-associated (abundance and/or capacity of WNV bridge vectors, host selection patterns, etc.) factors. Second, WNV human infections might be occurring but with a lower reporting fraction, which depends on factors that vary geographically within Europe, such as healthcare access, diagnostic awareness and surveillance practices. Third, since we had the objective to only map the bird-associated structural component of WNV infection risk, some areas might show high potential for wild bird communities to sustain a diffusion of WNV, although local vector population dynamics may not favour an active enzootic circulation yet (but may in the future). Indeed, in the definition of our structural risk metric, we could have accounted for the global spatial distribution of vector species that are key to WNV enzootic cycle. Nonetheless, contrary to other vector-borne diseases with a moving vector emergence front (e.g. *Aedes* mosquitoes for dengue in Europe or *Ixodes* ticks for Lyme disease in Canada [33]), some WNV vectors such as *C. pipiens* are native to Europe and are already widely spread across the continent [34]. Therefore, the heterogeneity in mosquito populations that matters to explain the spatial distribution of WNV infections in Europe, at the NUTS region scale, might be more conjectural (e.g. higher *Culex* abundance in some months because of climatic factors), thus out of the scope of our time-independent risk metric, than structural (global presence of a species in a region). Interestingly, WNV infections in birds of prey have been reported for the first time in 2024 in Latvia and Estonia [35,36], countries which globally display high BRI values but with no notified WNV human case as of October 2025 [26].

The BRI we computed may be used to strengthen WNV (active or passive) surveillance or sampling efforts in geographical areas that we estimated favourable to the virus’ enzootic circulation, despite the absence of case notification. Such initiatives may target the animal (e.g. wild birds, horses or zoo animals), human (e.g. serosurveys, or sensitisation of the general public or practitioners to WNV suggestive symptoms) and vector (e.g. viral testing of *Culex* mosquito pools) compartments, possibly as part of an integrated scheme [37,38].

Moreover, we compared the BRI, a time-aggregated measure of the structural risk, i.e. a zoonotic potential related to the structure of bird communities, to a time-aggregated West Nile disease outcome in humans. This is justified by the fact that the global spatial distribution of bird populations, while affected by the environmental crisis [39], is expected to change at the scale of several years or decades [3]. Because the occurrence of WNV outbreaks also depends on shorter term (e.g. monthly) variations of environmental variables and of the vector population, i.e. on the conjectural risk, the BRI should not be interpreted as a forecasting tool or a predictor of outbreak intensity. The next step will be to account for the mechanisms driving outbreaks at fine time scale in suitable areas [40], by implementing a One Health analysis framework combining bird, equine and human cases notification data, as [10]. The mixture of both the BRI and vector mapping would lead to a more accurate prediction of WNV infections at a finer spatio-temporal scale, in a silent circulation epidemiological context. This would also allow to assess the impact of shifts in vector populations related to climate change scenarios [41].

Although we extracted bird species’ spatial distributions from a recent study [25], other sources of data may provide maps of birds occurrence or abundance, such as the eBird Status and Trends dataset that is based on citizen science [19,20]. Provided that the 150 species we considered to build the BRI are available from eBird, it will be interesting in the future to compare the metric as computed from such alternative data sources.

Our Bird Risk Index increases with the number of bird species presenting life history traits associated to higher WNV seroprevalence. Consequently, it is partly driven by species richness, whose specific effect on disease occurrence has been debated in the literature as part of the ‘dilution effect’ hypothesis [42–44]. Furthermore, because seroprevalence is only an indicator of past exposure to the virus in surviving individuals, it is not sufficient to infer a bird species’ epidemiological role. Indeed, birds present varying competence for transmitting WNV to biting mosquitoes [45,46]. Hence a high seroprevalence may reflect a high exposure but not an important reservoir or amplification role. In contrast, the seroprevalence in surviving individuals might be low in a bird species with higher virus-induced lethality [45,47], even though it had an amplification role during an outbreak. For these reasons, the BRI may not be directly interpreted as a reservoir index, but rather as an index of potential exposure of a bird community to WNV, based on host traits. It should be noted that competence and lethality can be reliably estimated only as part of infection experiments, which are rare in European bird species, especially for passerines (e.g. [22,23]). For costs and ethical reasons, it is unlikely that such experiments will be conducted at large scale (e.g. in dozens of species) in the next years, maintaining uncertainty about these parameters in European bird populations. In this context, we focused on birds <50g because (i) their shorter lifespan [16,17] allows to relate more proportionally the seroprevalence to the virus exposure, and (ii) because they seem globally more abundant within bird communities than larger birds such as *Corvidae* and raptors (see Supplementary Figure S1).

Previous studies investigated traits factors associated with higher WNV seroprevalence in bird species. First, as already described [2,48], we confirmed that altricial birds tended to be more WNV seropositive than precocial birds, potentially due to lower feather coverage and higher exposure to mosquito bites in altricial chicks [49]. The latter mechanism may also explain the association we found for bird species nesting on the ground, as compared to nesting above ground [2]. Second, we showed a protective effect of bird gregariousness, similarly as a previous study [50]. This might be due to an encounter-dilution effect, a hypothesis which postulates that a higher number of host individuals in the group reduces the probability for each of them to be bitten by mosquito vectors and to be subsequently exposed to the virus [51]. Third, we did not identify a significant effect of migratory compared to non-migratory species, consistently with a study led in Senegal [50] but unlike other studies in Spain [52,53]. This point would need further investigations, since some bird migratory routes encompass West Africa and Europe, and were shown to favour WNV (re)introductions in both directions [54]. Furthermore, we did not distinguish the migratory species that winter in Europe and can breed north of our study area (e.g. *Calidris alpina*) from species that winter in Africa and breed in Europe (e.g. *Hirundo rustica*). However, whilst this means that WNV exposure may occur outside Europe [50], and that accounting for this multiple migration strategies may improve our Model 1, WNV infection of birds in Europe can theoretically occur across a quite large period (spring to autumn, depending on the regions) that encompass parts of the breeding and wintering seasons for many bird species. In the future, mechanistic models accounting for migratory and resident bird dynamics may help to explain the patterns observed in viral and serological data collected in bird communities. On top of long-distance migrations, traits related to birds’ local (within NUTS regions) movement abilities, such as home range size or natal dispersal distance, may drive the connectivity between bird populations and the diversity of land types and vectors encountered by individuals, thus affecting the exposure to WNV [55]. Therefore, not accounting for such traits when predicting WNV seroprevalence might bias our estimates of other traits’ effects in the case they are correlated, or lead to false biological interpretations for these effects (e.g. interpreting our results as a causal effect of trait A on WNV seroprevalence, whereas it is another unobserved trait B that impacts both trait A and WNV seroprevalence).

## Conclusion

To conclude, we computed a spatially heterogeneous metric quantifying WNV potential for circulation in European birds, which proved to be associated with WNV human case notification. It may be used to strengthen surveillance efforts in identified risk areas and, when associated with environmental and mosquito data, to better predict WNV outbreaks across Europe.

## Ethics statement

We only used data that was already published in the literature (serosurveys data) or publicly available data collected as part of routine surveillance (human cases data).

## Authors’ contributions

JB performed the literature review, retrieved and curated the data, carried out the formal model analysis and wrote the first draft of the manuscript. JB, RM and BD designed the study, interpreted the results, reviewed and edited the manuscript.

## Data availability statement

The data and code reproducing this study are available from https://doi.org/10.5281/zenodo.18163635. The human cases data is publicly available from the European Centre for Disease Prevention and Control website: https://www.ecdc.europa.eu/en/infectious-disease-topics/west-nile-virus-infection/surveillance-and-updates-west-nile-virus.

