## Supplementary materials for "Mapping the Bird Risk Index for West Nile virus in Europe and its relationship with disease occurrence in humans"

**Supplementary Data 1** (attached file). Life history traits associated to 150 European bird species weighting less than 50g (initially published by [1]).

**Supplementary Data 2** (attached file). References and data included in the model predicting WNV seroprevalence in bird species (Model 1).

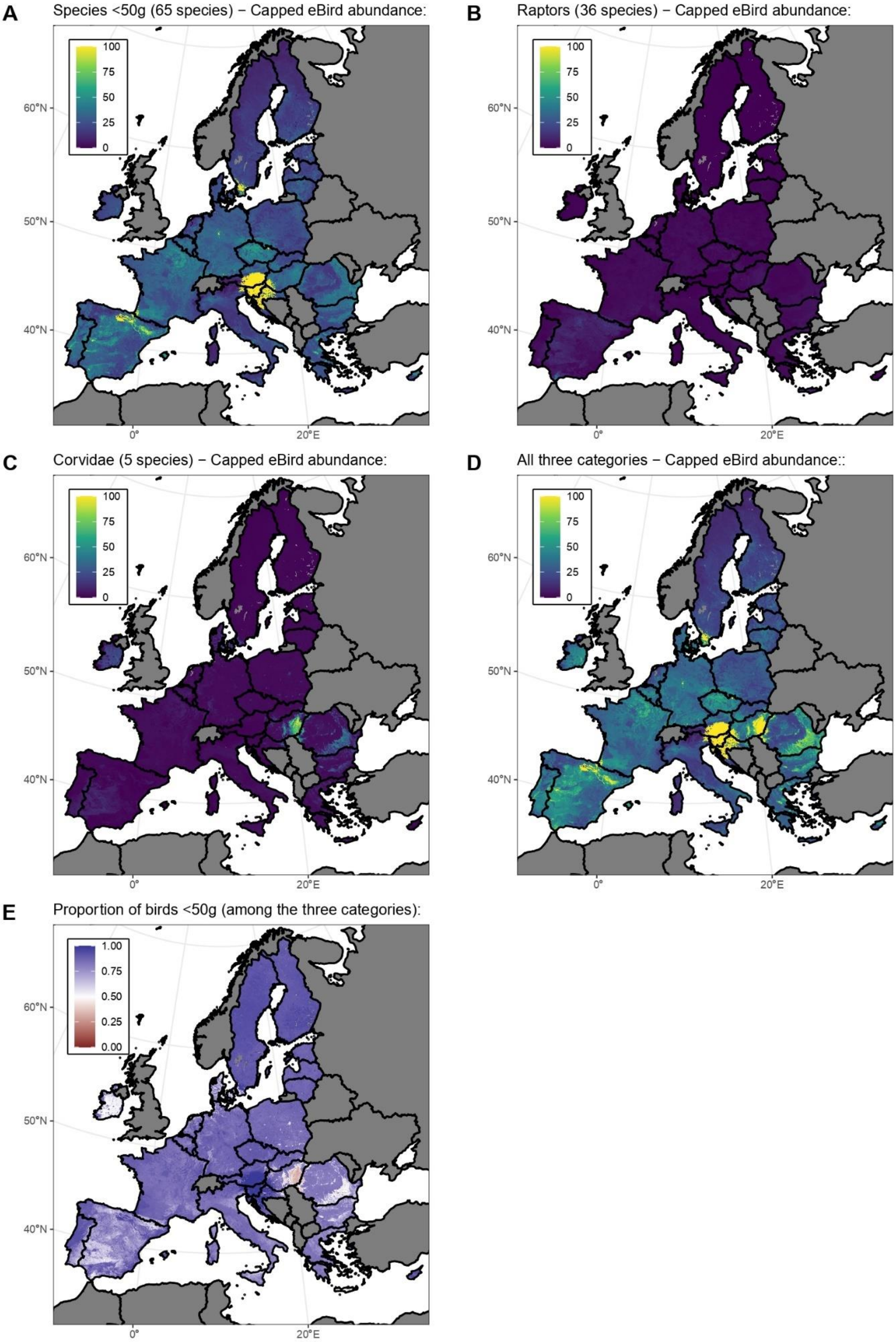
A
Species <50g (65 species) − Capped eBird abundance:
B
Raptors (36 species) − Capped eBird abundance:
C
Corvidae (5 species) − Capped eBird abundance:
D
All three categories − Capped eBird abundance::
E
Proportion of birds <50g (among the three categories):
100
75
50
25
0
1.00
0.75
0.50
0.25
0.00
60°N
50°N
40°N
0°
20°E

**Supplementary Figure S1.** Cumulated relative abundance of European species of birds weighting less than 50g (panel A), of raptors (panel B), of *Corvidae* (panel C) and of the three categories combined (panel D), as provided in the 2023 eBird Status and Trends dataset [2,3]. Relative abundance is defined as the expected probability of encountering a species, for an expert eBird observer as part of a 2 km, 1 hour travel at the optimal time of the day and for optimal weather conditions, multiplied by the expected count of the species conditional on its occurrence at the given location. For non-resident species, we considered the average of the relative abundances across seasons. The color scale is capped at 100 to facilitate visualization. Panel E depicts the proportion of birds <50g among all three categories. This dataset only contains a subset of all species of these three categories: 65 species of European birds <50g (*Prunella collaris, Phylloscopus borealis, Riparia riparia, Hirundo rustica, Sylvia atricapilla, Phoenicurus ochruros, Acrocephalus dumetorum, Luscinia svecica, Cyanistes caeruleus, Fringilla montifringilla, Cettia cetti, Emberiza cirlus, Locustella naevia, Fringilla coelebs, Phylloscopus collybita, Alcedo atthis, Luscinia megarhynchos, Phoenicurus phoenicurus, Actitis hypoleucos, Apus apus, Galerida cristata, Calidris alpina, Prunella modularis, Ficedula hypoleuca, Erithacus rubecula, Serinus serinus, Acrocephalus scirpaceus, Jynx torquilla, Passer montanus, Regulus regulus, Parus major, Acrocephalus arundinaceus, Motacilla cinerea, Eremophila alpestris, Passer domesticus, Phylloscopus ibericus, Calcarius lapponicus, Lanius minor, Calidris minuta, Anthus pratensis, Oenanthe oenanthe, Emberiza hortulana, Apus pallidus, Lanius collurio, Loxia curvirostra, Phalaropus lobatus, Anthus petrosus, Monticola saxatilis, Cercotrichas galactotes, Locustella luscinioides, Certhia brachydactyla, Alauda arvensis, Plectrophenax nivalis, Calidris temminckii, Galerida theklae, Luscinia luscinia, Anthus trivialis, Phylloscopus bonelli, Motacilla alba, Loxia leucoptera, Phylloscopus trochilus, Lanius senator, Emberiza aureola, Emberiza citrinella, Cisticola juncidis*), 36 species of raptors (*Terathopius ecaudatus, Elanus caeruleus, Milvus migrans, Aquila fasciata, Hieraaetus pennatus, Aegolius funereus, Aegypius monachus, Buteo buteo, Neophron percnopterus, Falco eleonorae, Pernis apivorus, Gyps fulvus, Falco tinnunculus, Accipiter nisus, Aquila chrysaetos, Strix nebulosa, Gypaetus barbatus, Clanga pomarina, Falco naumanni, Athene noctua, Asio otus, Falco columbarius, Circus pygargus, Surnia ulula, Circus cyaneus, Pernis ptilorhynchus, Pandion haliaetus, Falco peregrinus, Milvus milvus, Buteo lagopus, Asio flammeus, Circaetus gallicus, Aquila adalberti, Aquila nipalensis, Strix aluco, Gyps africanus*) and 5 species of *Corvidae* (*Cyanopica cooki, Corvus corax, Garrulus glandarius, Pica pica, Corvus frugilegus*).

**Supplementary Table S1.** Eco-epidemiological hypotheses that justify the inclusion of variables (life history traits) in Model 1.

| Variable | Eco-epidemiological hypothesis |
|---|---|
| Migratory status (yes or no) | Migratory birds are exposed to enzootic WNV circulation when they spend part of the year in sub-saharian areas [4]. |
| Body mass (under 50g) | Larger birds release more $CO_2$, hence may attract mosquito-vectors more. |
| Nest height (ground, intermediate or above 4 meters) | Nest height may affect the density of mosquitoes around the nest [5], and therefore the exposure of individual birds to mosquito-vectors [1]. |
| Use of urban/suburban habitat (yes or no) | The type of habitat affects the community of hosts (including of non-avian hosts) and vectors [6], and therefore may impact the rate of bites of individual birds by infected mosquitoes. |
| Nocturnal gregariousness (yes or no) | Birds that gather in groups may release more $CO_2$ and attract mosquitoes more than solitary individuals [7]. On the contrary, in groups of birds, an encounter-dilution effect may decrease the exposure of each individual bird to mosquito bites, due to an increase of the bird-to-mosquito ratio [8]. |
| Exposure of nestlings (altricial or precocial) | Altricial nestlings are immobile, have little defensive behavior against mosquitoes and are not protected by feather coverage, which exposes them more to their bites [1,7,9]. |
| Breeding sociality (yes or no) | Birds that gather in groups may release more $CO_2$ and attract mosquitoes more than solitary individuals [7]. On the contrary, in groups of birds, an encounter-dilution effect may decrease the exposure of each individual bird to mosquito bites, due to an increase of the bird-to-mosquito ratio [8]. |

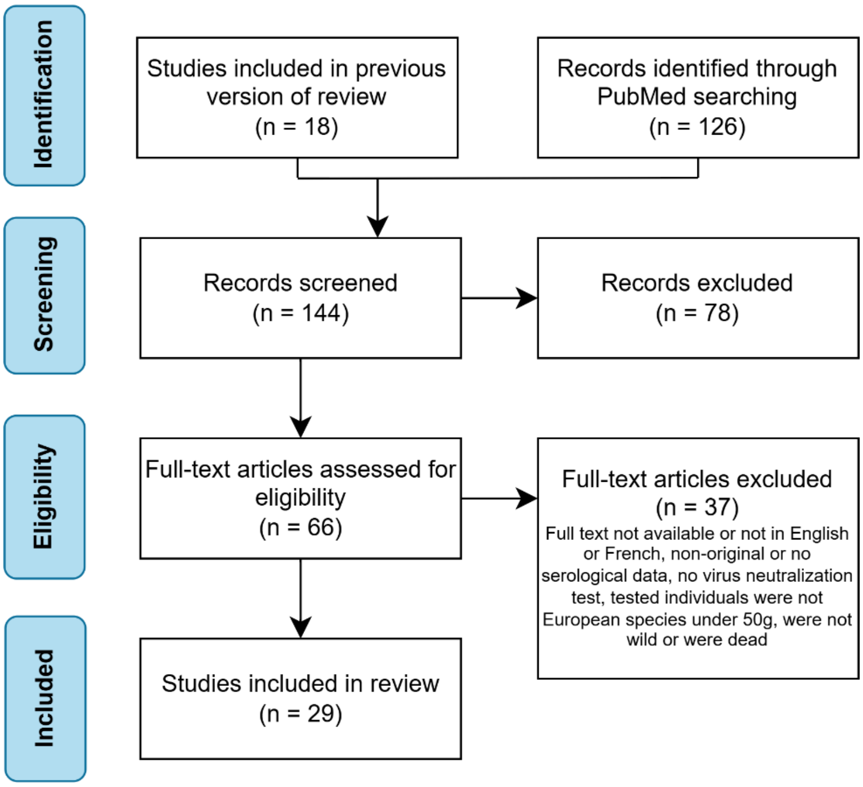


**Supplementary Figure S2.** Flow diagram of the literature review. Twenty-nine studies were included in the review [4,10–37].

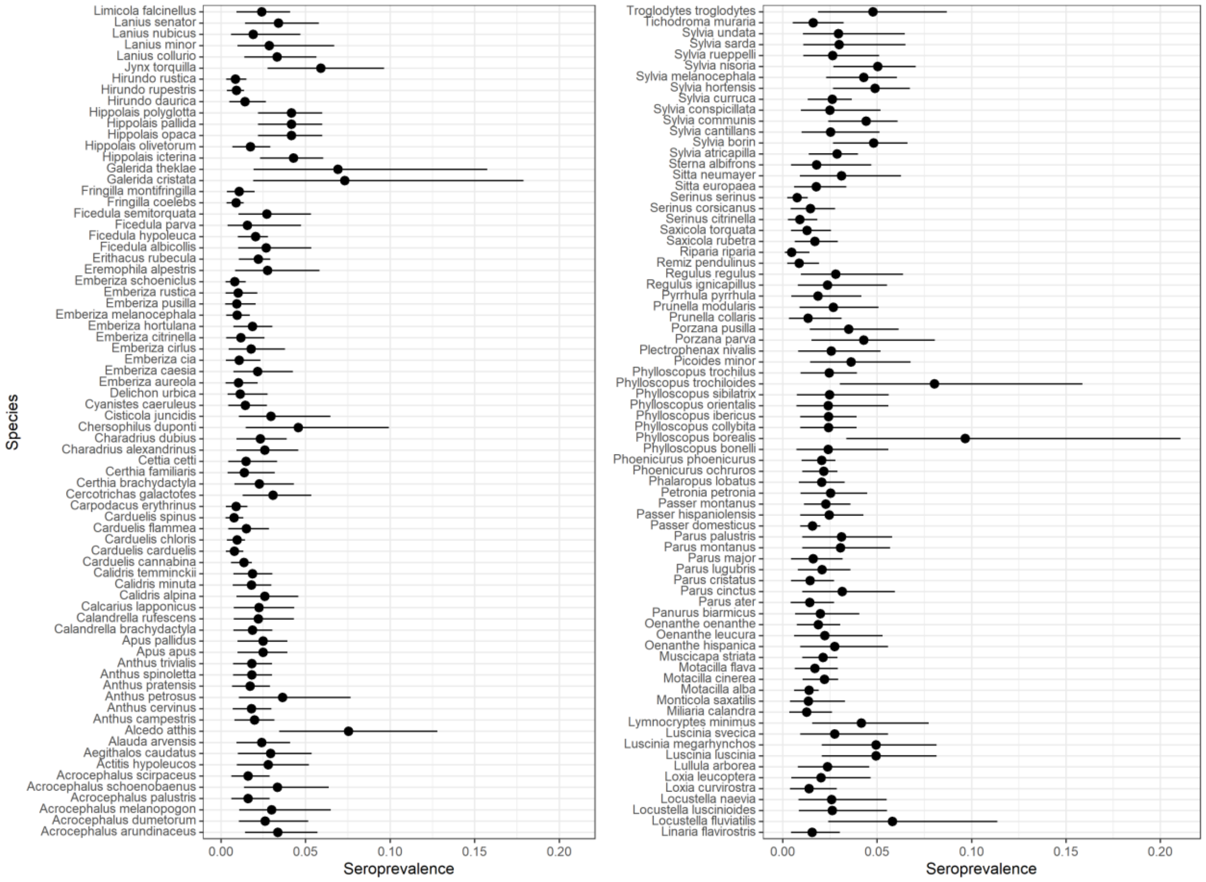


**Supplementary Figure S3.** WNV seroprevalence in 150 European bird species predicted by Model 1. For each species, the predicted point value is when all the literature data was included in the model, and the interval corresponds to the 95% confidence interval using 1,000 bootstrap replicates.

**Supplementary Table S2.** Results of the 5-fold cross-validation of Model 2. For each cross-validation fold, we fitted the model to the training dataset, resulting in posterior estimates of the fixed effects coefficients, and of hyperparameters of the BYM2 (precision $\tau_{BYM2}$ and mixing parameter $\phi_{BYM2}$) and "Country" (precision $\tau_{Country}$) random effects. We then compared model predictions to the testing data, by computing the deviance-based overall (or conditional) $R^2$, where the null model has no fixed (other than the intercept) or random effect, and marginal $R^2$, where the null model has an intercept and the BYM2 and "Country" random effects. For the final model, random effects' hyperparameters ($\tau_{BYM2}$, $\phi_{BYM2}$ and $\tau_{Country}$) were estimated at respectively 0.62 [0.49; 0.82], 0.26 [0.12; 0.48] and 0.093 [0.034; 0.24].

| **Iteration** | **Posterior estimates of fixed effects coefficients (95% credible interval)** | | | **Hyperparameters of the BYM2 and Country effects (95% credible interval)** | | | **Validation metrics** | |
|---|---|---|---|---|---|---|---|---|
| | Intercept | Mean BRI | log(population density) | $\tau_{BYM2}$ | $\phi_{BYM2}$ | $\tau_{Country}$ | Overall $R^2$ | Marginal $R^2$ |
| 1 | -23.6 [-28.46; -19.14] | 9.64 [7.21; 12.16] | 0.97 [0.69; 1.25] | 0.59 [0.44; 0.83] | 0.45 [0.08; 0.87] | 0.09 [0.038; 0.206] | 0.64 | 0.37 |
| 2 | -24.62 [-29.77; -19.98] | 10.48 [8.05; 13.07] | 1.02 [0.73; 1.31] | 0.68 [0.51; 0.94] | 0.43 [0.2; 0.71] | 0.084 [0.026; 0.23] | 0.5 | 0.21 |
| 3 | -24.51 [-29.54; -19.84] | 9.73 [7.24; 12.31] | 1.05 [0.75; 1.36] | 0.53 [0.4; 0.72] | 0.44 [0.16; 0.74] | 0.097 [0.032; 0.267] | 0.67 | 0.34 |
| 4 | -25.43 [-30.14; -20.99] | 9.48 [7.16; 11.86] | 1.14 [0.86; 1.43] | 0.67 [0.49; 0.95] | 0.26 [0.05; 0.78] | 0.097 [0.039; 0.226] | 0.56 | 0.27 |
| 5 | -24.32 [-29.06; -19.97] | 10.27 [7.92; 12.72] | 1.03 [0.76; 1.32] | 0.72 [0.52; 1.03] | 0.34 [0.06; 0.83] | 0.103 [0.04; 0.22] | 0.5 | 0.22 |

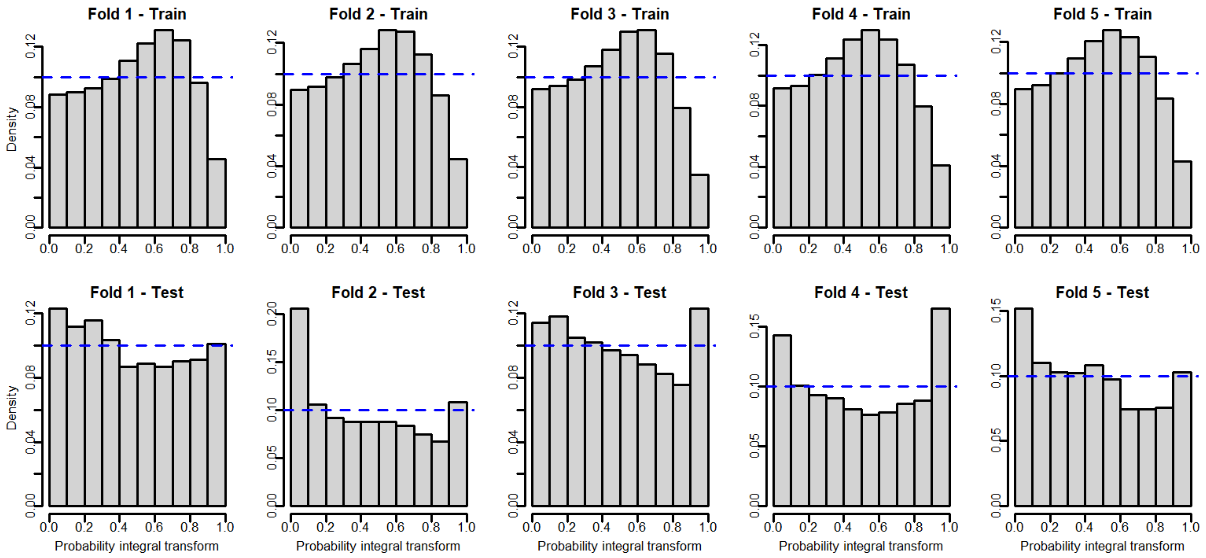


**Supplementary Figure S4.** Probability Integral Transform (PIT) histograms for Model 2, assessing the consistency between model predictions and the training (first row) and testing (second row) data, for each cross-validation fold (columns 1 to 5).

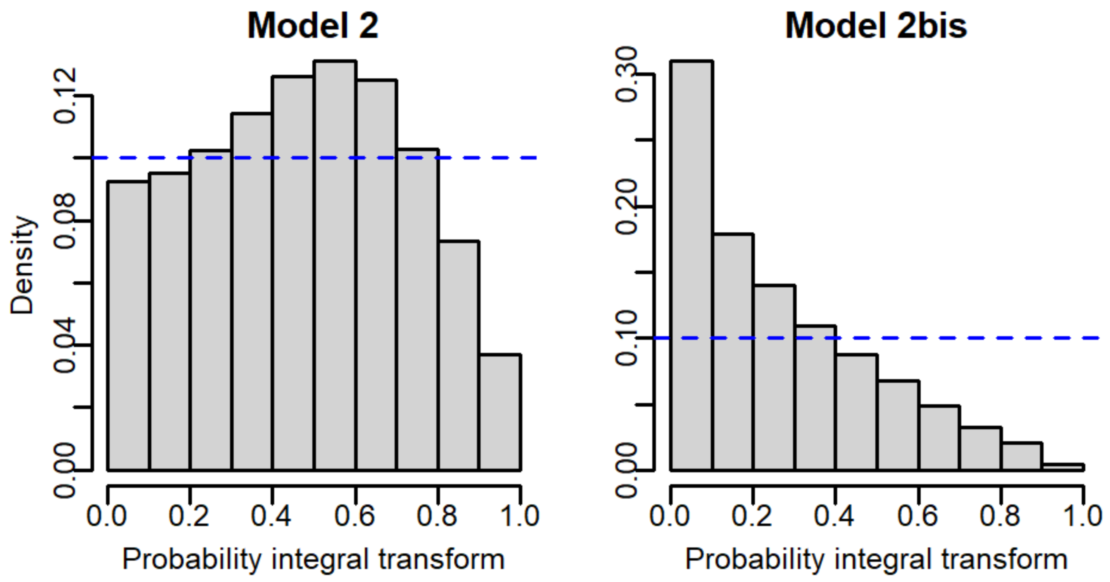


**Supplementary Figure S5.** Probability Integral Transform (PIT) histograms for Models 2 (left panel) and 2bis (right panel), assessing the consistency between final model predictions and the full dataset.

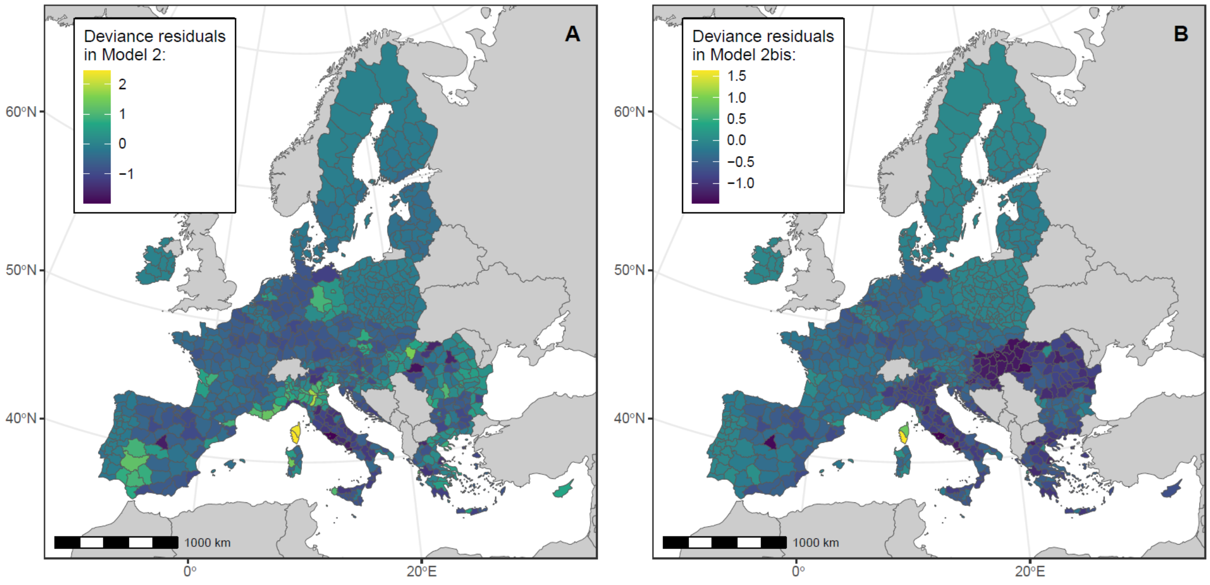


**Supplementary Figure S6.** Deviance residuals in Model 2 and 2bis, predicting respectively the number of years with notified WNV human cases and the cumulated number of notified WNV human cases, in NUTS European administrative regions between 2016 and 2023. We performed analyses at the NUTS3 level, or NUTS2 level for Belgium, Germany, Malta and Netherlands.

**Supplementary Table S3.** Sensitivity analysis for Model 2: for each alternative model tested, the included predictors, value of the Deviance Information Criterion (DIC) and coefficient posterior estimates (with their 95% credible interval) are shown. The baseline model used in the main analysis is on the first row.

| Predictors included | DIC | Parameter | Posterior estimate |
|---|---|---|---|
| Intercept + Mean BRI + log(population) + random effects | 1156.0 | Intercept | -24.7 [-29.2; -20.6] |
| | | Mean BRI | 9.97 [7.82; 12.2] |
| | | log(population) | 1.06 [0.80; 1.32] |
| Intercept only + random effects | 1207.5 | Intercept | -5.51 [-7.29; -4.25] |
| Intercept only | 2791.4 | Intercept | -2.09 [-2.17; -2.0] |
| Intercept + log(population) + random effects | 1234.6 | Intercept | -19.6 [-24.0; -15.5] |
| | | log(population) | 1.10 [0.81; 1.40] |
| Intercept + Species richness + log(population) + random effects | 1167.5 | Intercept | -23.7 [-28.3; -19.7] |
| | | Species richness | 8.82 [6.86; 10.9] |
| | | log(population) | 1.04 [0.78; 1.31] |
| Intercept + P50 + log(population) + random effects | 1190.9 | Intercept | -21.4 [-25.5; -17.6] |
| | | P50 | 3.75 [2.94; 4.59] |
| | | log(population) | 1.06 [0.81; 1.32] |

*BRI: Bird Risk Index; P50: proportion of bootstrap replicates for which each pixel fell in the 50% highest pixel values.*

**Supplementary Table S4.** Results of Model 2bis: the coefficient posterior estimates (with their 95% credible interval) are shown.

| Predictors included | Parameter | Posterior estimate |
|---|---|---|
| Intercept + Mean BRI + offset(log(population)) + random effects | (Intercept) | -21.43 [-25.27; -18.31] |
| | PIOU_moyen | 9.63 [6.42; 13.27] |

*BRI: Bird Risk Index.*

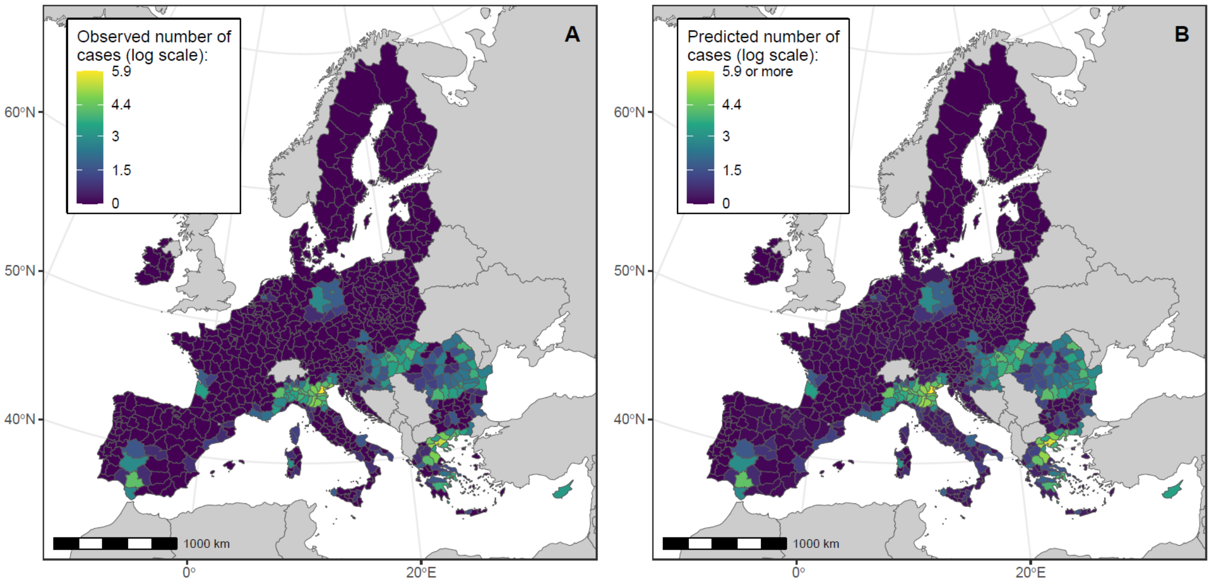


**Supplementary Figure S7.** Model 2bis maps showing the observed (panel A) and predicted (panel B) cumulated number of notified WNV human cases in NUTS European administrative regions between 2016 and 2023, on the log scale. We performed analyses at the NUTS3 level, or NUTS2 level for Belgium, Germany, Malta and Netherlands.

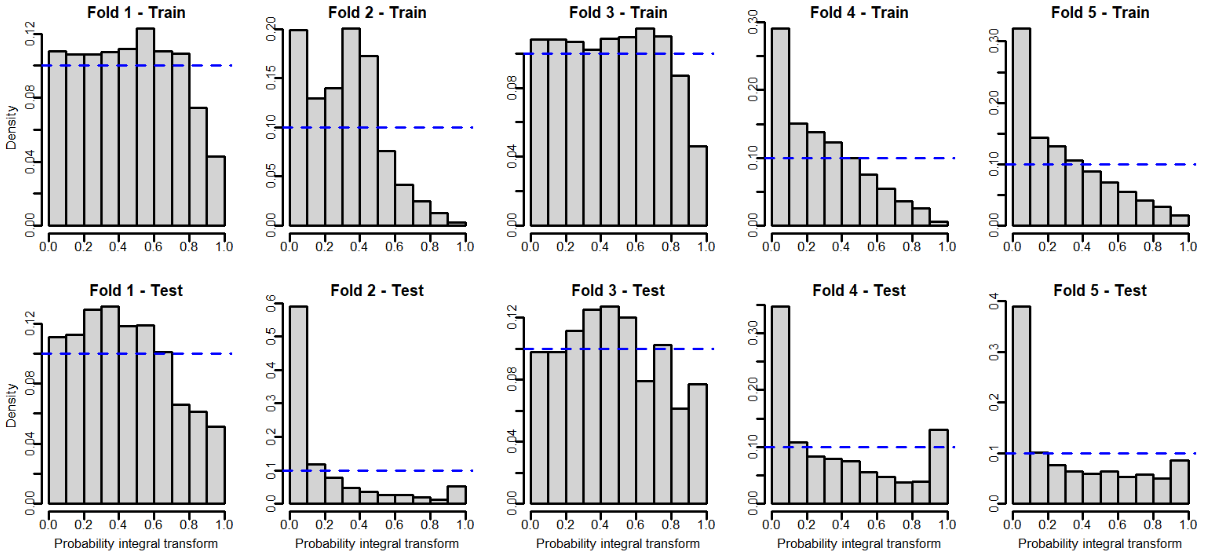


**Supplementary Figure S8.** Probability Integral Transform (PIT) histograms for Model 2bis, assessing the consistency between model predictions and the training (first row) and testing (second row) data, for each cross-validation fold (columns 1 to 5).
